# Adaptive short term COVID-19 prediction for India

**DOI:** 10.1101/2020.07.18.20156745

**Authors:** Shuvrangshu Jana, Debasish Ghose

## Abstract

In this paper, a data-driven adaptive model for infection of COVID-19 is formulated to predict the confirmed total cases and active cases of an area over 4 weeks. The parameter of the model is always updated based on daily observations. It is found that the short term prediction of up to 3-4 weeks can be possible with good accuracy. Detailed analysis of predicted value and the actual value of confirmed total cases and active cases for India from 1^st^ June to 3^rd^ July is provided. Prediction over 7, 14, 21, 28 days has the accuracy about 0.73% ± 1.97%, 1.92% ± 2.95%, 4.34% ± 3.91%, 6.40% ± 9.26% of the actual value of confirmed total cases. Similarly, the 7, 14, 21, 28 days prediction has the accuracy about 1.24% ± 6.57%, 3.04% ± 10.00%, 6.33% ± 16.12%, 10.20% ± 24.14% of the actual value of confirmed active cases.

## 1. Introduction

The accurate predictions of the spread of the novel coronavirus (COVID-19) are essential for planning and management of medical resources as well as lockdown strategy. A good prediction can avoid the shortage of critical medical resources, economical loss associated with unnecessary lockdowns. Mathematical modelling of infectious disease is generally performed by classifying the population into different compartments such as Susceptible, Infectious, Recovered, Deceased. Various classical compartmental model reported in literature such as SIR, SIS, SIRD, MSIR, SEIR, SEIS, MSEIR, MSEIRS. In order to model the spreading of corona virus, various modification of the SIR model is proposed in literature to include travel information [1], behavioural changes [2], various intervention measures such as quarantine, strict social distancing, tracing and isolation [3],[4], [5]. The model parameters are generally obtained for a specific area as there can be huge variation in these parameters based on the government policies, economical factors, adoption of social distancing measures. Prediction for the Spread of COVID-19 for a specific country is developed such for India [6, 7], China [8, 9, 10], South Korea-Italy [1], Iran [11],Korea [2], UK [12].

The estimation of spread of COVID-19 is made based on the assumption on rate of growth of total cumulative cases following a certain pattern such as gaussian [13], Verhulst equation [14], Lotka-Volterra dynamic model[15], Gompertz equation [16]. Estimation for Greece, Netherlands, Germany, Italy, Spain, France, United Kingdom and United States is developed using Gaussian fitting hypothesis based on the assumptions that evolutions of infected cases are of Gaussian in nature [13]. In [17], estimations for COVID-19 is performed from the fatality data for India under the assumption that the growth of infected cases is exponential. In [15], Lotka-Volterra dynamic model is used to represent the growth rate and the model coefficients are derived using the Extended Kalman filter techniques. In [18], Kalman filter-based techniques are used for short term prediction model.

Time-invariant SIR model is not effective to predict the spread of corona virus[19], considering the fact the large variation in the transmission dynamics. Also, it is difficult to fit a single model in different areas. So, Prediction models are developed based on the time series analysis of the reported data, where the model parameters c are estimated based on the previous observations of the confirmed cases. Data-driven estimation methods using long short-term memory (LSTM) and curve fitting is used for prediction of COVID-19 cases for India in 30 days ahead [20]. Deceased cases of COVID-19 prediction for one month is developed using a hybrid model using discrete wavelet decomposition and autoregressive integrated moving average (ARIMA) models [21]. In [22], Autoregressive Integrated moving average model is used to predict the daily number of COVID-9 cases up to 4 weeks. A mathematical model for prediction is developed using power series polynomials, where the coefficients of the polynomials are obtained using the least square approximations.

Generally, it is observed that it is difficult to predict in the long term due to the various uncertainties involved in the government decision-making process, social distancing measured followed by the people, availability of medical resources. There is a time lag between the time of infection and the time of reporting of infection. So in short term prediction has less uncertainty compared to the long term. A good general prediction model should use information about the local state as less as possible.

In this paper, a short term prediction model is proposed based on the concept of SIR model, where the parameters are updated continuously based on the observations using weighted least square techniques. It is observed that the spread of COVID-19, recovery and deceased rate can be approximated using time-varying polynomials. The proposed model is validated by comparing the true and actual value of June for India over the past predictions on time different time windows such as 7 days, 14 days, 21 days, and 28 days. In case of 2 days time window, It is found that error in prediction is within 6.40 % ± 9.26 % and 10.22 % ± 24.14 % for the prediction of total cases and active cases.

The rest of the paper is described as follows. The basic model is described in Section 2. The model parameters are estimated in Section 3. The proposed model is validated with COVID-19 statistics of India in Section 5. Expected total and active cases for a month are predicted in Section 7. The prediction results are discussed and concluded in Discussions and Conclusions Sections respectively.

## 2. Model formulation

The total number of people who have been infected with the virus can be classified as active, recovered or deceased cases. Few cases will be unreported due to non-development of symptoms in the infected person. In Fig. 1, different categories of different cases are shown. We will consider the population as susceptible (*S*), Infectious (*I*), and Removed (as shown in Fig. 2). Infectious cases are divided into two categories, active cases(*A*) and active unreported cases *A*_ur_. Similarly, recovered and deceased cases are also classified as those who are reported and unreported cases. So, if *N* is the total population,then,

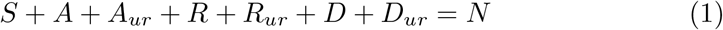

**Figure 1:**
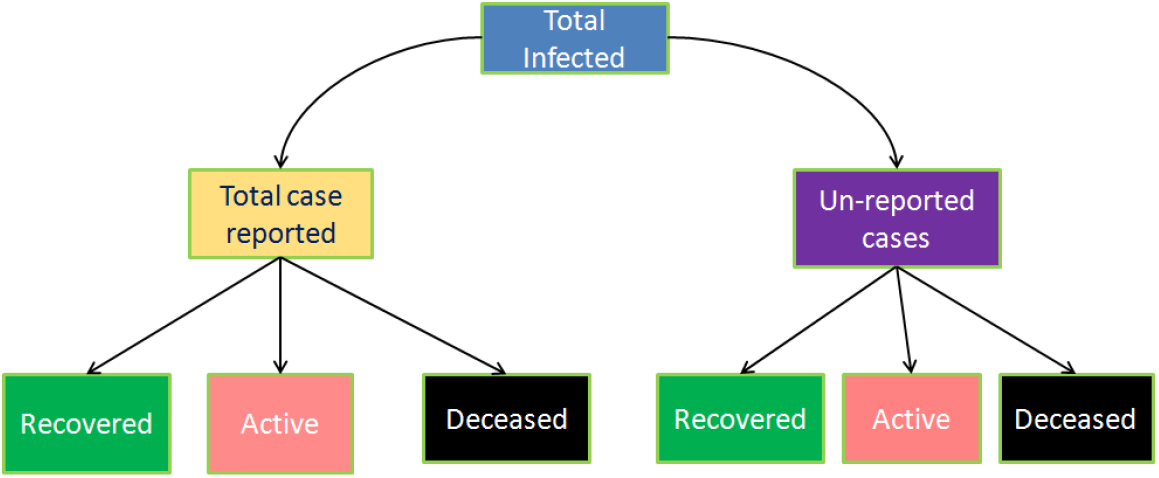

**Figure 2:**
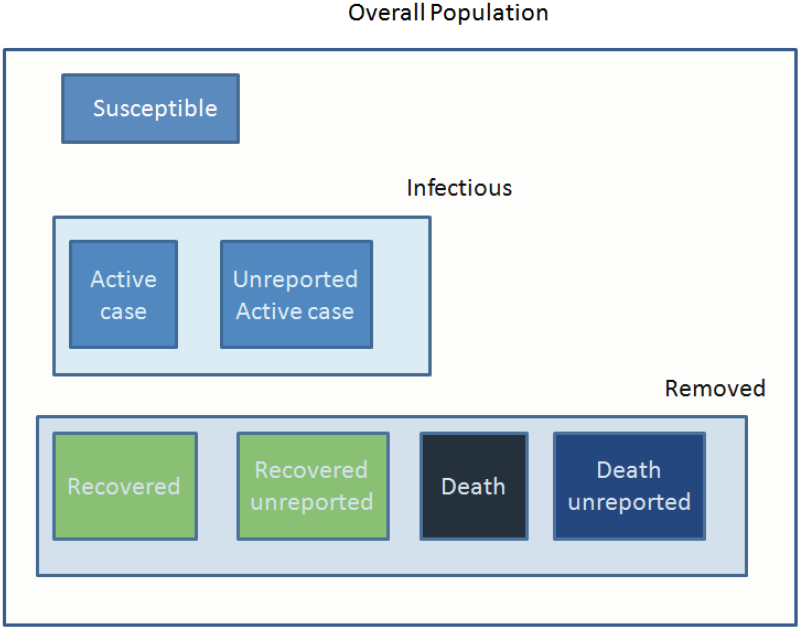
Different compartment

where, *R, R*_ur_ are the reported recovered and unreported recovered cases respectively. Similarly, *D, D*_ur_ are the reported and unreported deceased cases respectively. Also,

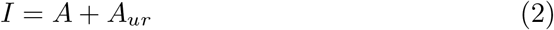

As per standard SIR model, If *β* is the infection rate, then,

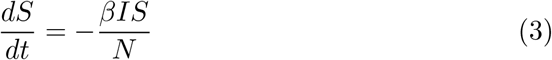

Also, we can write,

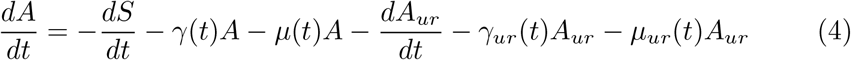

where *γ*(*t*) and *µ*(*t*) are the rate of recovered and death among the reported cases; and *γ*_ur_ and *µ*_ur_ are the rate of recovered and death cases among unreported cases.

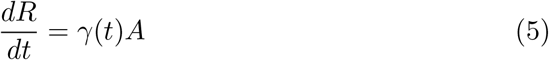

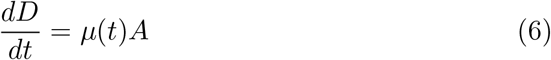

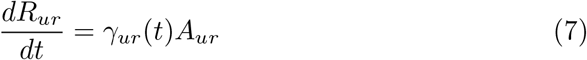

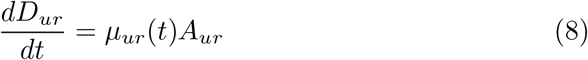

## 3. Parameter estimation

The different parameter of the above model is estimated from the observed data and dynamically adjusted using the daily observations. The active cases, recovered and deceased from the reported cases are readily available; whereas, the similar statistics for the unreported cases need to be estimated from random antibody tests or serological tests. The confirmed active cases at each day are obtained by the following equation

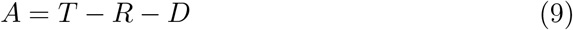

where, T is the total number of reported infected person. Then we have,

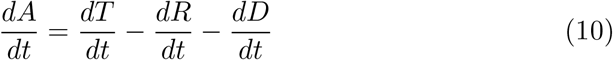

From the previous reported daily statistics of daily new infected cases, recovered cases, and deceased cases; the rate of growth of the total number of cases, recovered cases and deceased cases are obtained.

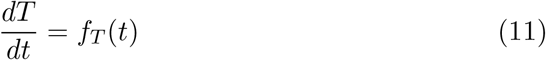

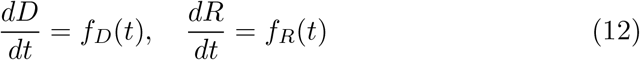

The rate of growth of total cases are obtained by taking derivative of the variable T. T can be represented by different functions. In the case of India, it is observed that T can be approximated using a fourth-order polynomial. Let consider T be,

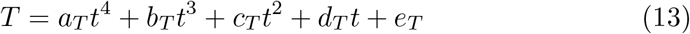

The coefficients (*a*_*T*_, *b*_*T*_, *c*_*T*_, *d*_*T*_, *e*_*T*_) of the function T is obtained using the weighted least squares method by minimizing the error between the predicted value and the observed daily values. The weighted least squares method is used to provide more weightage on the recent values which in turn reflects the status of lockdown, social distancing measured followed by the citizens. The following cost function is used.

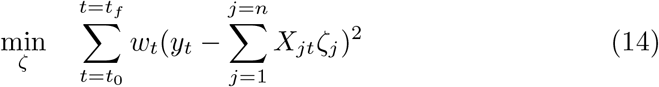

where *y*_t_ is the observed cumulative total cases, *T* is expressed as *T* = *Xζ*, and *ζ* are the coefficients of the polynomial. The weights (*w*_t_ are selected from the previous observations as 1(last 7 days), 0.9 (last 7-14 days), 0.8 (last 14-21 days), 0.5 (last 21-28 days) and 0.2 for further previous days.

The weights need to be selected properly for an area considering the lockdown/unlock period. The optimum values of *ζ* are calculated from the equation 14 as follows.

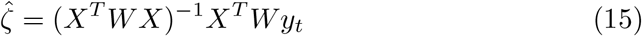

where *W* is the diagonal matrix consist of weights (*w*_t_). It is observed that reported recovered and deceased cases can be approximated using the fourth-order and second-order polynomial respectively.

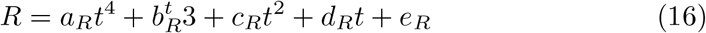

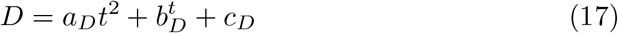

## 4. Prediction

Prediction values are updated based on the current observations. The data of total cases recovered cases, and deceased cases from 14^th^ March to 3^rd^ July for India is considered in the analysis. The data is collected from the “https://api.covid19india.org/” website, where the daily COVID-19 updates of various states and central government’s of India are recorded. The variation of total cases, cumulative recovered cases, cumulative deceased cases, and corresponding active cases on daily basis are plotted in Figure 3a, Figure 3b, Figure 3c, Figure 3d respectively. Based on the observations till 3^rd^ July, the prediction for the next 14 days is shown in Figure 4a and Figure 4b. The variation of coefficients of the prediction curve of the total cases is tabulated in Table 1.

**Table 1:** Total cases prediction curve coefficients

| **Date of prediction** | $a_T$ | $b_T$ | $c_T$ | $d_T$ | $e_T$ |
| --- | --- | --- | --- | --- | --- |
| 03.07.2020 | 0.0031 | 0.0442 | 8.85 | -116.96 | 595.4 |
| 02.07.2020 | 0.003 | 0.0496 | 8.59 | -112.71 | 580.6 |
| 01.07.2020 | 0.003 | 0.0548 | 8.38 | -110.48 | 578.6 |
| 30.06.2020 | 0.0028 | 0.0889 | 6.34 | -68.53 | 381.5 |
| 29.06.2020 | 0.0025 | 0.1615 | 1.86 | 26.12 | -72.7 |
| 28.06.2020 | 0.002 | 0.2576 | -4.13 | 153.64 | -687.4 |
| 27.06.2020 | 0.0016 | 0.3317 | -8.71 | 250.11 | -1148.3 |
| 26.06.2020 | 0.0013 | 0.3811 | -11.67 | 311.14 | -1433.9 |
| 25.06.2020 | 0.0012 | 0.4033 | -12.94 | 336.07 | -1546.1 |
| 24.06.2020 | 0.0012 | 0.4038 | -12.88 | 333.34 | -1528.1 |
| 23.06.2020 | 0.012 | 0.3984 | -12.47 | 323.59 | -1478 |
| 22.06.2020 | 0.001 | 0.4402 | -14.92 | 372.64 | -1701.1 |
| 21.06.2020 | 0.0006 | 0.5134 | -19.29 | 461.18 | -2106.9 |
| 20.06.2020 | 0.0004 | 0.5559 | -21.85 | 513.52 | -2347.7 |
| 19.06.2020 | 0.0004 | 0.5502 | -21.60 | 509.79 | -2335.1 |
| 18.06.2020 | 0.0007 | 0.5007 | -18.86 | 457.595 | -2108.5 |
| 17.06.2020 | 0.0012 | 0.4045 | -13.50 | 355.37 | -1665 |
| 16.06.2020 | 0.0019 | 0.2981 | -7.59 | 243.34 | -1181.7 |
| 15.06.2020 | 0.0022 | 0.2338 | -4.21 | 181.97 | -925.9 |
| 14.06.2020 | 0.0024 | 0.2027 | -2.61 | 153.62 | -810.5 |
| 13.06.2020 | 0.0027 | 0.153 | 0.0002 | 106.73 | -617.8 |
| 12.06.2020 | 0.0031 | 0.0952 | 3.0 | 53.13 | -399.5 |
| 11.06.2020 | 0.0036 | 0.0046 | 7.72 | -30.60 | -59.9 |
| 10.06.2020 | 0.0042 | -0.0945 | 12.85 | -121.40 | 306 |
| 9.06.2020 | 0.0044 | -0.12 | 14.08 | -143.07 | 393.2 |
| 8.06.2020 | 0.0043 | -0.11 | 13.43 | -131.40 | 345.8 |
| 7.06.2020 | 0.0044 | -0.116 | 13.92 | -139.45 | 376.6 |
| 6.06.2020 | 0.0044 | -0.119 | 14.06 | -141.79 | 385.7 |
| 5.06.2020 | 0.0045 | -0.13 | 14.67 | -152.24 | 425.9 |
| 4.06.2020 | 0.0047 | -0.16 | 16.17 | -176.80 | 518.1 |
| 3.06.2020 | 0.005 | -0.204 | 18.14 | -208.85 | 637.4 |
| 2.06.2020 | 0.0047 | -0.169 | 16.43 | -180.86 | 533.3 |
| 1.06.2020 | 0.0044 | -0.12 | 14.19 | -144.43 | 399 |

**Figure 3:**
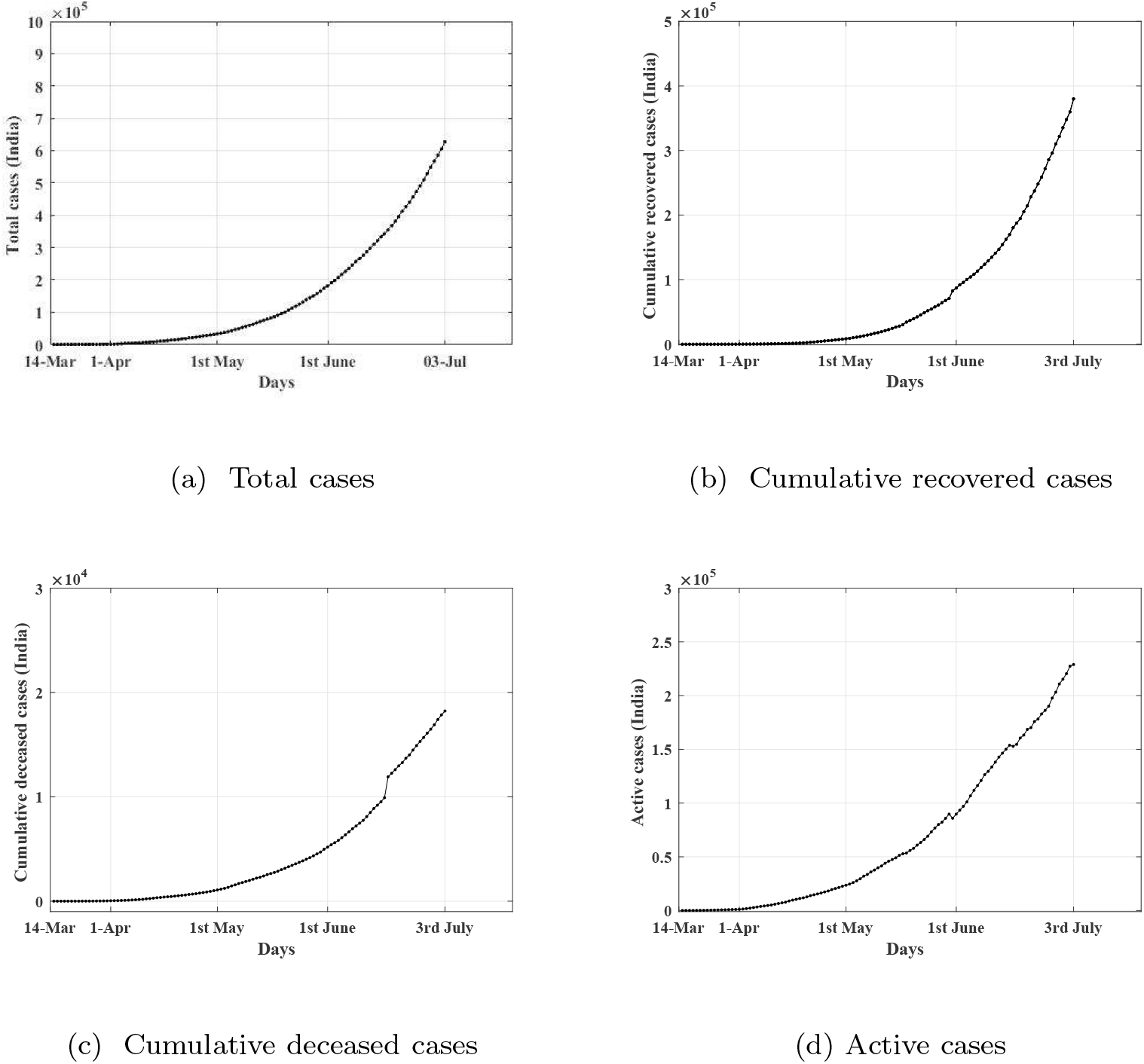
Total case, Recovered case, Deceased case, Active Case

**Figure 4:**
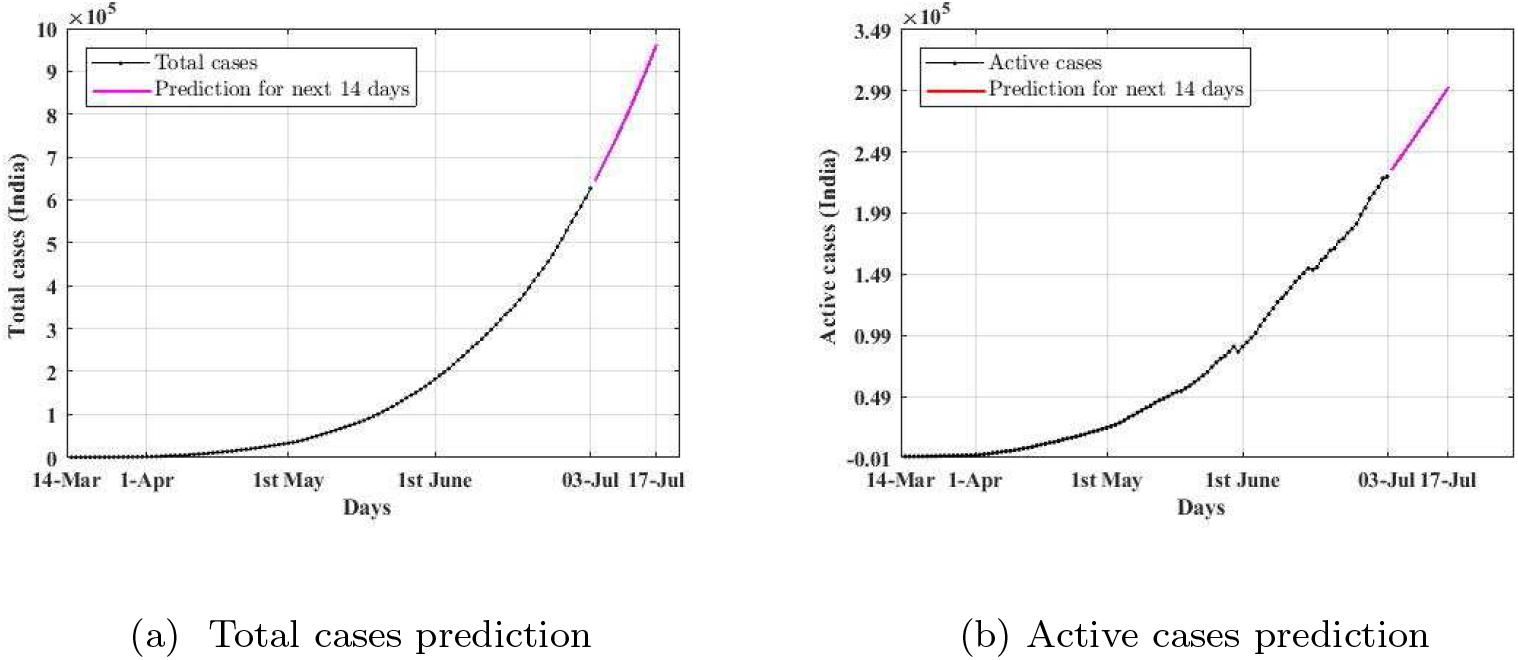
Prediction of total cases and active cases

## 5. Model Validation

### 5.1. Total case prediction validation

The difference between the predicted value and the actual value is compared to check the efficiency of the prediction algorithm. The predicted value on the 7^th^, 14^th^, 21^th^ and 28^th^ days back is considered for comparison. The predictions from June 1^st^ to July 3^rd^ are considered for detailed analysis. The total case prediction over a 7 days time window and the corresponding actual value is shown in Table 2. In 26^th^ June, the total cases prediction for 3^rd^ July was 61130 and the actual value reported is 627065. So, the error in prediction is 2.54 % of the actual value of 627065. From Table 2, over the complete duration of June 1^st^ to July 3^rd^, the error between the actual and predicted value is 0.73 % ± 1.97 % of the actual value.

**Table 2:** Validation of prediction for 7 days window

| Date during pre-<br>diction | Date of prediction | Prediction | Actual | Error from<br>actual(%) |
| --- | --- | --- | --- | --- |
| 26.06.2020 | 03.07.2020 | 611130 | 627065 | -2.54 |
| 25.06.2020 | 02.07.2020 | 590940 | 605118 | -2.27 |
| 24.06.2020 | 01.07.2020 | 571970 | 585689 | -2.31 |
| 23.06.2020 | 30.06.2020 | 553680 | 567433 | -2.26 |
| 22.06.2020 | 29.06.2020 | 534770 | 549094 | -2.47 |
| 21.06.2020 | 28.06.2020 | 516140 | 529484 | -2.40 |
| 20.06.2020 | 27.06.2020 | 499230 | 509342 | -1.91 |
| 19.06.2020 | 26.06.2020 | 484050 | 491087 | -1.30 |
| 18.06.2020 | 25.06.2020 | 470350 | 472882 | -0.58 |
| 17.06.2020 | 24.06.2020 | 457990 | 456014 | +0.40 |
| 16.06.2020 | 23.06.2020 | 445710 | 440358 | +1.25 |
| 15.06.2020 | 22.06.2020 | 432190 | 426798 | +1.62 |
| 14.06.2020 | 21.06.2020 | 418110 | 411647 | +1.87 |
| 13.06.2020 | 20.06.2020 | 404670 | 395729 | +2.44 |
| 12.06.2020 | 19.06.2020 | 391710 | 381091 | +2.94 |
| 11.06.2020 | 18.06.2020 | 379170 | 367262 | +3.33 |
| 10.06.2020 | 17.06.2020 | 367390 | 354154 | +3.76 |
| 9.06.2020 | 16.06.2020 | 355690 | 343068 | +3.67 |
| 8.06.2020 | 15.06.2020 | 342430 | 333036 | +3.01 |
| 7.06.2020 | 14.06.2020 | 329090 | 321631 | +2.55 |
| 6.06.2020 | 13.06.2020 | 316490 | 309592 | +2.43 |
| 5.06.2020 | 12.06.2020 | 304000 | 298286 | +2.17 |
| 4.06.2020 | 11.06.2020 | 292000 | 287151 | +1.89 |
| 3.06.2020 | 10.06.2020 | 280810 | 275995 | +1.53 |
| 2.06.2020 | 9.06.2020 | 270090 | 266014 | +1.31 |
| 1.06.2020 | 8.06.2020 | 258580 | 257478 | + 0.77 |
| 31.05.2020 | 7.06.2020 | 247430 | 246596 | +0.33 |
| 30.05.2020 | 6.06.2020 | 237260 | 236188 | +0.25 |
| 29.05.2020 | 5.06.2020<sub>1</sub> | 228180 | 226716 | +0.62 |
| 28.05.2020 | 4.06.2020 | 219640 | 216869 | +0.13 |
| 27.05.2020 | 3.06.2020 | 211570 | 207180 | +1.90 |
| 26.05.2020 | 2.06.2020 | 202590 | 198368 | +1.95 |
| 25.05.2020 | 1.06.2020 | 192820 | 190645 | +1.20 |

The predicted and actual values over a 14 days prediction window from June 1^st^ is tabulated in Table 3. As per Table 3 In case of prediction on 14 days duration, the error between the actual and the predicted value is found to be 1.92 % ± 2.95 % of the actual value. Similary, prediction over 21 days and 28 days time window are tabulated in 4 and 5. The error between the predicted value and the actual value over 21 days and 28 days is 4.34 % ± 3.91 % and 6.40 % ± 9.26 % of the actual value respectively.

**Table 3:** Validation of prediction for 14 days window

| Date during pre-<br>diction | Date of prediction | Prediction | Actual | Error from<br>actual(%) |
| --- | --- | --- | --- | --- |
| 19.06.2020 | 03.07.2020 | 603000 | 627065 | -3.84 |
| 18.06.2020 | 02.07.2020 | 588280 | 605118 | -2.78 |
| 17.06.2020 | 01.07.2020 | 576050 | 585689 | -1.65 |
| 16.06.2020 | 30.06.2020 | 563900 | 567433 | -0.62 |
| 15.06.2020 | 29.06.2020 | 549090 | 549094 | -0.0007 |
| 14.06.2020 | 28.06.2020 | 532640 | 529484 | +0.60 |
| 13.06.2020 | 27.06.2020 | 517310 | 509342 | +1.56 |
| 12.06.2020 | 26.06.2020 | 502820 | 491087 | +2.39 |
| 11.06.2020 | 25.06.2020 | 488880 | 472882 | +3.38 |
| 10.06.2020 | 24.06.2020 | 476370 | 456014 | +4.46 |
| 9.06.2020 | 23.06.2020 | 463910 | 440358 | +5.35 |
| 8.06.2020 | 22.06.2020 | 448000 | 426798 | +4.97 |
| 7.06.2020 | 21.06.2020 | 431400 | 411647 | +4.80 |
| 6.06.2020 | 20.06.2020 | 416030 | 395729 | +5.13 |
| 5.06.2020 | 19.06.2020 | 400600 | 381091 | +5.12 |
| 4.06.2020 | 18.06.2020 | 385890 | 367262 | +5.07 |
| 3.06.2020 | 17.06.2020 | 372490 | 354154 | +5.18 |
| 2.06.2020 | 16.06.2020 | 359780 | 343068 | +4.87 |
| 1.06.2020 | 15.06.2020 | 344840 | 333036 | +3.54 |
| 31.05.2020 | 14.06.2020 | 330210 | 321631 | +2.67 |
| 30.05.2020 | 13.06.2020 | 317490 | 309592 | +2.55 |
| 29.05.2020 | 12.06.2020 | 306910 | 298286 | +2.89 |
| 28.05.2020 | 11.06.2020 | 297380 | 287151 | +3.56 |
| 27.05.2020 | 10.06.2020 | 288690 | 275995 | +4.60 |
| 26.05.2020 | 9.06.2020 | 277750 | 266014 | +4.41 |
| 25.05.2020 | 8.06.2020 | 264890 | 257478 | +2.88 |
| 24.05.2020 | 7.06.2020 | 251330 | 246596 | +1.92 |
| 23.05.2020 | 6.06.2020 | 238310 | 236188 | +0.89 |
| 22.05.2020 | 5.06.2020<sub>12</sub> | 226270 | 226716 | -0.20 |
| 21.05.2020 | 4.06.2020 | 214030 | 216869 | -1.31 |
| 20.05.2020 | 3.06.2020 | 201940 | 207180 | -2.53 |
| 19.05.2020 | 2.06.2020 | 192680 | 198368 | -2.87 |
| 18.05.2020 | 1.06.2020 | 183700 | 190645 | -3.64 |

**Table 4:** Validation of prediction for 21 days window

| Date during pre-<br>diction | Date of prediction | Prediction | Actual | Error from<br>actual(%) |
| --- | --- | --- | --- | --- |
| 12.06.2020 | 03.07.2020 | 636020 | 627065 | +1.43 |
| 11.06.2020 | 02.07.2020 | 621080 | 605118 | +2.64 |
| 10.06.2020 | 01.07.2020 | 608540 | 585689 | +3.90 |
| 9.06.2020 | 30.06.2020 | 596090 | 567433 | +5.05 |
| 8.06.2020 | 29.06.2020 | 577340 | 549094 | +5.14 |
| 7.06.2020 | 28.06.2020 | 556870 | 529484 | +5.17 |
| 6.06.2020 | 27.06.2020 | 538410 | 509342 | +5.71 |
| 5.06.2020 | 26.06.2020 | 519600 | 491087 | +5.81 |
| 4.06.2020 | 25.06.2020 | 501850 | 472882 | +6.13 |
| 3.06.2020 | 24.06.2020 | 486130 | 456014 | +6.60 |
| 2.06.2020 | 23.06.2020 | 471440 | 440358 | +7.06 |
| 1.06.2020 | 22.06.2020 | 452240 | 426798 | + 5.96 |
| 31.05.2020 | 21.06.2020 | 433210 | 411647 | + 5.24 |
| 30.05.2020 | 20.06.2020 | 417510 | 395729 | +5.50 |
| 29.05.2020 | 19.06.2020 | 405600 | 381091 | + 6.43 |
| 28.05.2020 | 18.06.2020 | 395580 | 367262 | +7.71 |
| 27.05.2020 | 17.06.2020 | 386990 | 354154 | +9.27 |
| 26.05.2020 | 16.06.2020 | 374040 | 343068 | +9.03 |
| 25.05.2020 | 15.06.2020 | 357380 | 333036 | +7.31 |
| 24.05.2020 | 14.06.2020 | 338890 | 321631 | +5.37 |
| 23.05.2020 | 13.06.2020 | 321070 | 309592 | +3.71 |
| 22.05.2020 | 12.06.2020 | 304740 | 298286 | +2.16 |
| 21.05.2020 | 11.06.2020 | 287530 | 287151 | -0.13 |
| 20.05.2020 | 10.06.2020 | 270180 | 275995 | -2.11 |
| 19.05.2020 | 9.06.2020 | 257870 | 266014 | -3.06 |
| 18.05.2020 | 8.06.2020 | 245800 | 257478 | -4.53 |
| 17.05.2020 | 7.06.2020 | 236170 | 246596 | -4.23 |
| 16.05.2020 | 6.06.2020 | 234360 | 236188 | -0.77 |
| 15.05.2020 | 5.06.2020<sub>13</sub> | 231570 | 226716 | -2.14 |
| 14.05.2020 | 4.06.2020 | 228380 | 216869 | +5.31 |
| 13.05.2020 | 3.06.2020 | 224320 | 207180 | +8.27 |
| 12.05.2020 | 2.06.2020 | 218790 | 198368 | +10.30 |
| 11.05.2020 | 1.06.2020 | 208670 | 190645 | +9.45 |

**Table 5:** Validation of prediction for 28 days window

| Date during pre-<br>diction | Date of prediction | Prediction | Actual | Error from<br>actual(%) |
| --- | --- | --- | --- | --- |
| 5.06.2020 | 03.07.2020 | 664400 | 627065 | +5.95 |
| 4.06.2020 | 02.07.2020 | 643270 | 605118 | +6.30 |
| 3.06.2020 | 01.07.2020 | 625210 | 585689 | +6.75 |
| 2.06.2020 | 30.06.2020 | 608650 | 567433 | +7.26 |
| 1.06.2020 | 29.06.2020 | 584200 | 549094 | +6.39 |
| 31.05.2020 | 28.06.2020 | 559650 | 529484 | +5.69 |
| 30.05.2020 | 27.06.2020 | 540520 | 509342 | +6.12 |
| 29.05.2020 | 26.06.2020 | 527610 | 491087 | +7.44 |
| 28.05.2020 | 25.06.2020 | 517820 | 472882 | +9.50 |
| 27.05.2020 | 24.06.2020 | 510370 | 456014 | +11.92 |
| 26.05.2020 | 23.06.2020 | 495480 | 440358 | +12.52 |
| 25.06.2020 | 22.06.2020 | 474220 | 426798 | +11.11 |
| 24.05.2020 | 21.06.2020 | 449360 | 411647 | +9.16 |
| 23.05.2020 | 20.06.2020 | 425280 | 395729 | +7.47 |
| 22.05.2020 | 19.06.2020 | 403440 | 381091 | +5.86 |
| 21.05.2020 | 18.06.2020 | 379570 | 367262 | +3.35 |
| 20.05.2020 | 17.06.2020 | 355050 | 354154 | -0.25 |
| 19.05.2020 | 16.06.2020 | 338820 | 343068 | -1.24 |
| 18.05.2020 | 15.06.2020 | 322730 | 332036 | -3.09 |
| 17.05.2020 | 14.06.2020 | 310660 | 321631 | -2.89 |
| 16.05.2020 | 13.06.2020 | 312340 | 309592 | +0.88 |
| 15.05.2020 | 12.06.2020 | 312310 | 298286 | +4.45 |
| 14.05.2020 | 11.06.2020 | 311560 | 287151 | +8.19 |
| 13.05.2020 | 10.06.2020 | 309330 | 275995 | +12.07 |
| 12.05.2020 | 9.06.2020 | 304560 | 266014 | +14.49 |
| 11.05.2020 | 8.06.2020 | 291510 | 257478 | +13.22 |
| 10.05.2020 | 7.06.2020 | 291040 | 246596 | +18.02 |
| 9.05.2020 | 6.06.2020 | 287750 | 236188 | +21.83 |
| 8.05.2020 | 5.06.2020<sub>14</sub> | 274110 | 226716 | +20.90 |
| 7.05.2020 | 4.06.2020 | 250330 | 216869 | +15.43 |
| 6.05.2020 | 3.06.2020 | 219070 | 207180 | +5.74 |
| 5.05.2020 | 2.06.2020 | 174510 | 198368 | -12.03 |
| 4.05.2020 | 1.06.2020 | 137740 | 190645 | -27.75 |

The error in prediction from mid-April to July 3^rd^ is plotted in Figure 5a. In Figure5a, magenta points show the predicted value on 7 days back and the black points are the predicted value on that day. At each day, the difference between the black and magenta point is the difference in predicted and the actual value. Similar plots for the prediction window of 14 days, 21 days, and 28 days are plotted in the Figure 5b, Figure 5c, and Figure 5d respectively.

**Figure 5:**
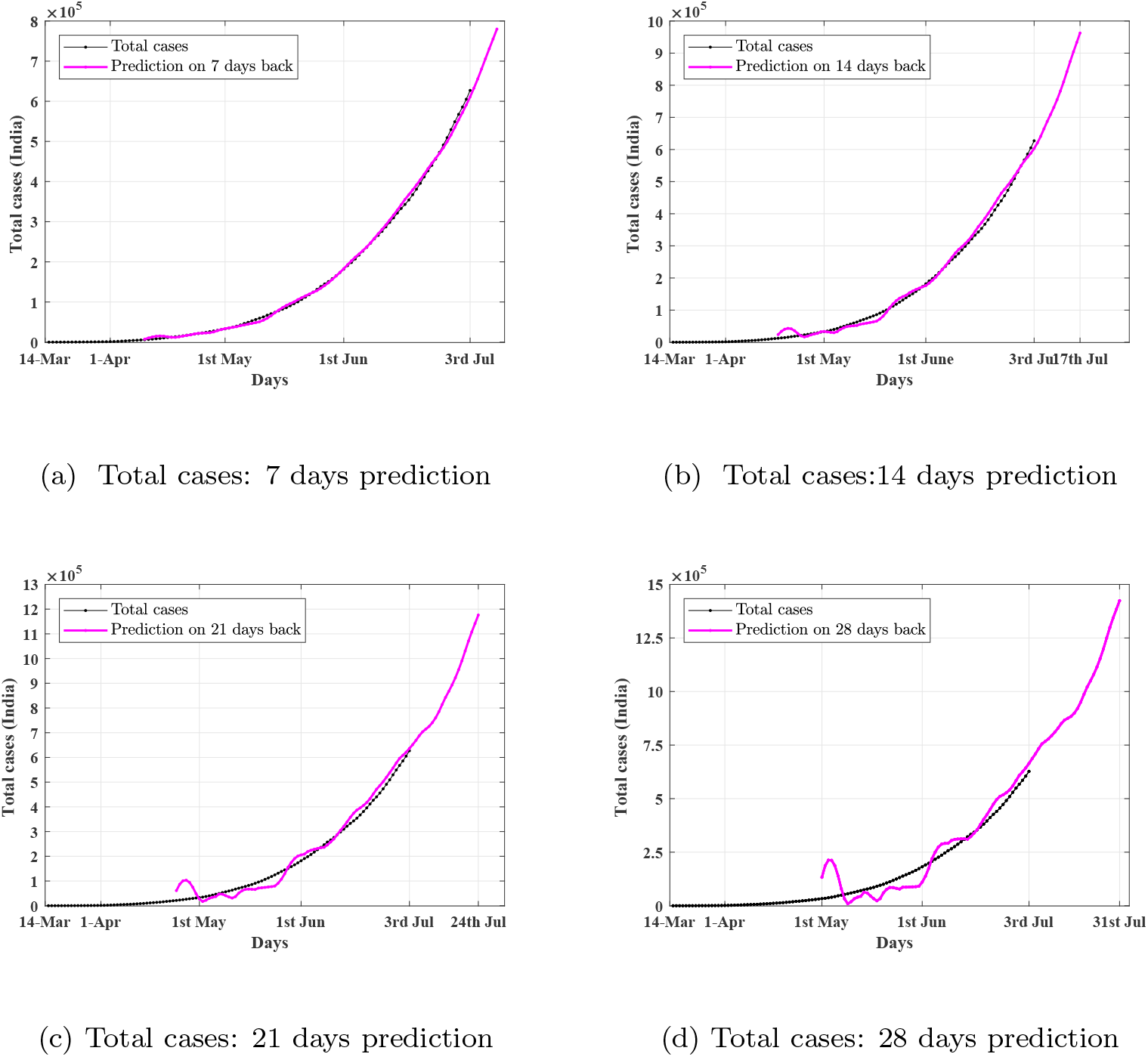
Validation of prediction of total cases

### 5.2. Active case prediction validation

The prediction results in case of active cases are compared with the actual value of the active cases. The prediction values and the error in prediction of active cases over 7 days, 14 days, 21 days, 28 days time duration are tabulated in Table 6, Table 7, Table 8, and Table 9 respectively. The difference between the predicted active case and the actual active case is found to be 1.24 % ± 6.57 %, 3.04 % ± 10.00 %, 6.33 % ± 16.12 %, 10.20 % ± 24.14 % for the prediction window of 7 days, 14 days, 21 days, 28 days respectively. The error in active case prediction from mid April to July 3^rd^ over different time window is plotted in Figure 6a, Figure 6b, Figure 6c, and Figure 6d.

**Table 6:**
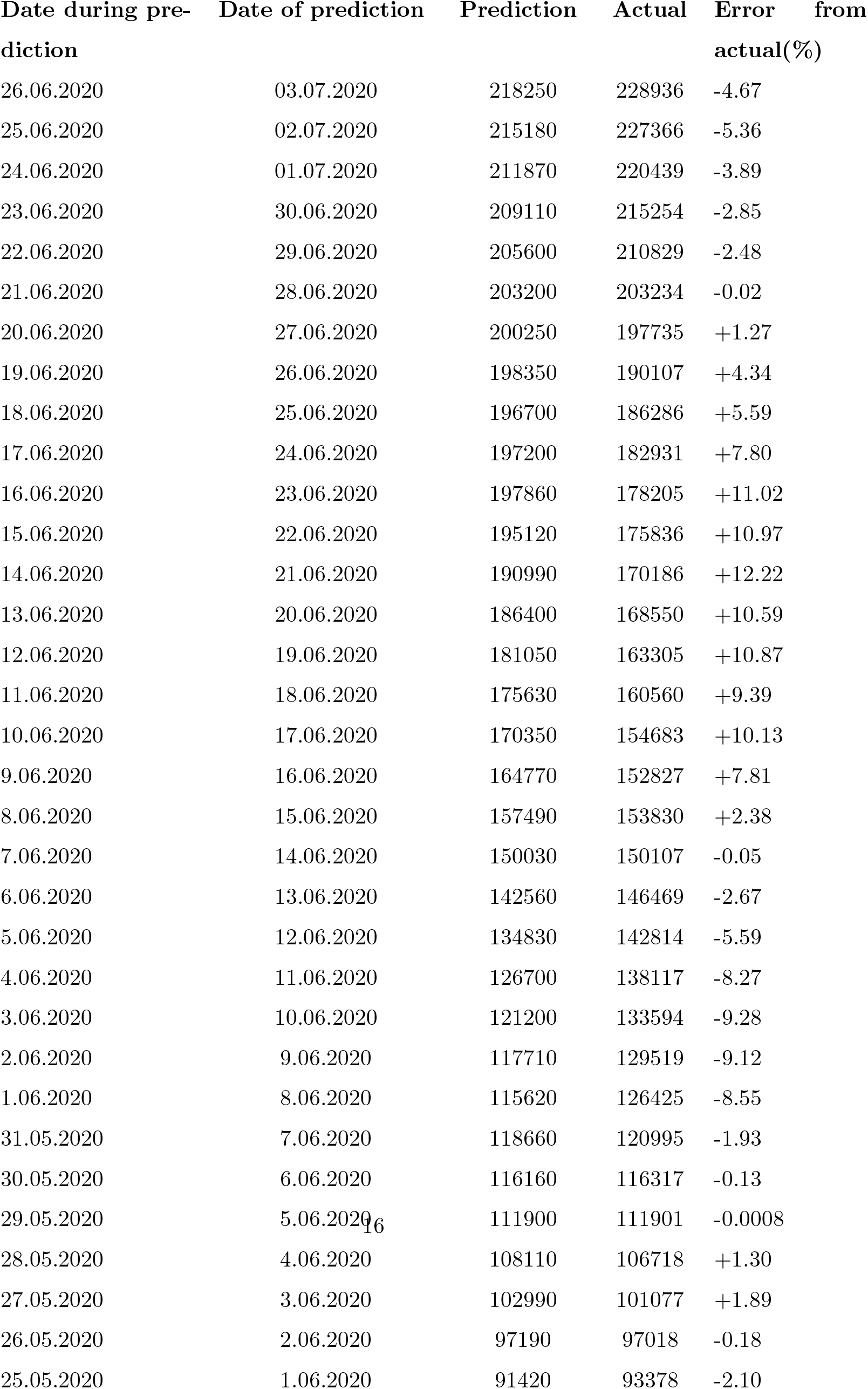
Validation of active case prediction for 7 days window

| Date during pre-<br>diction | Date of prediction | Prediction | Actual | Error from<br>actual(%) |
| --- | --- | --- | --- | --- |
| 26.06.2020 | 03.07.2020 | 218250 | 228936 | -4.67 |
| 25.06.2020 | 02.07.2020 | 215180 | 227366 | -5.36 |
| 24.06.2020 | 01.07.2020 | 211870 | 220439 | -3.89 |
| 23.06.2020 | 30.06.2020 | 209110 | 215254 | -2.85 |
| 22.06.2020 | 29.06.2020 | 205600 | 210829 | -2.48 |
| 21.06.2020 | 28.06.2020 | 203200 | 203234 | -0.02 |
| 20.06.2020 | 27.06.2020 | 200250 | 197735 | +1.27 |
| 19.06.2020 | 26.06.2020 | 198350 | 190107 | +4.34 |
| 18.06.2020 | 25.06.2020 | 196700 | 186286 | +5.59 |
| 17.06.2020 | 24.06.2020 | 197200 | 182931 | +7.80 |
| 16.06.2020 | 23.06.2020 | 197860 | 178205 | +11.02 |
| 15.06.2020 | 22.06.2020 | 195120 | 175836 | +10.97 |
| 14.06.2020 | 21.06.2020 | 190990 | 170186 | +12.22 |
| 13.06.2020 | 20.06.2020 | 186400 | 168550 | +10.59 |
| 12.06.2020 | 19.06.2020 | 181050 | 163305 | +10.87 |
| 11.06.2020 | 18.06.2020 | 175630 | 160560 | +9.39 |
| 10.06.2020 | 17.06.2020 | 170350 | 154683 | +10.13 |
| 9.06.2020 | 16.06.2020 | 164770 | 152827 | +7.81 |
| 8.06.2020 | 15.06.2020 | 157490 | 153830 | +2.38 |
| 7.06.2020 | 14.06.2020 | 150030 | 150107 | -0.05 |
| 6.06.2020 | 13.06.2020 | 142560 | 146469 | -2.67 |
| 5.06.2020 | 12.06.2020 | 134830 | 142814 | -5.59 |
| 4.06.2020 | 11.06.2020 | 126700 | 138117 | -8.27 |
| 3.06.2020 | 10.06.2020 | 121200 | 133594 | -9.28 |
| 2.06.2020 | 9.06.2020 | 117710 | 129519 | -9.12 |
| 1.06.2020 | 8.06.2020 | 115620 | 126425 | -8.55 |
| 31.05.2020 | 7.06.2020 | 118660 | 120995 | -1.93 |
| 30.05.2020 | 6.06.2020 | 116160 | 116317 | -0.13 |
| 29.05.2020 | 5.06.2020<sup>16</sup> | 111900 | 111901 | -0.0008 |
| 28.05.2020 | 4.06.2020 | 108110 | 106718 | +1.30 |
| 27.05.2020 | 3.06.2020 | 102990 | 101077 | +1.89 |
| 26.05.2020 | 2.06.2020 | 97190 | 97018 | -0.18 |
| 25.05.2020 | 1.06.2020 | 91420 | 93378 | -2.10 |

**Table 7:** Validation of active case prediction for 14 days window

| Date during pre-<br>diction | Date of prediction | Prediction | Actual | Error from<br>actual(%) |
| --- | --- | --- | --- | --- |
| 19.06.2020 | 03.07.2020 | 231130 | 228936 | + 0.96 |
| 18.06.2020 | 02.07.2020 | 231470 | 227366 | +1.81 |
| 17.06.2020 | 01.07.2020 | 236290 | 220439 | +7.19 |
| 16.06.2020 | 30.06.2020 | 241850 | 215254 | +12.36 |
| 15.06.2020 | 29.06.2020 | 240990 | 210829 | +14.31 |
| 14.06.2020 | 28.06.2020 | 237250 | 203234 | +16.74 |
| 13.06.2020 | 27.06.2020 | 232780 | 197735 | +17.72 |
| 12.06.2020 | 26.06.2020 | 226850 | 190107 | +19.33 |
| 11.06.2020 | 25.06.2020 | 220720 | 186286 | +18.48 |
| 10.06.2020 | 24.06.2020 | 214960 | 182931 | +17.51 |
| 9.06.2020 | 23.06.2020 | 208780 | 178205 | +17.16 |
| 8.06.2020 | 22.06.2020 | 199000 | 175836 | +13.17 |
| 7.06.2020 | 21.06.2020 | 188500 | 170186 | +10.76 |
| 6.06.2020 | 20.06.2020 | 177690 | 168550 | +5.42 |
| 5.06.2020 | 19.06.2020 | 166080 | 163305 | -1.70 |
| 4.06.2020 | 18.06.2020 | 151960 | 160560 | -5.36 |
| 3.06.2020 | 17.06.2020 | 143240 | 154683 | -7.40 |
| 2.06.2020 | 16.06.2020 | 138830 | 152827 | -9.16 |
| 1.06.2020 | 15.06.2020 | 137290 | 153830 | -10.75 |
| 31.05.2020 | 14.06.2020 | 140460 | 150107 | -6.43 |
| 30.05.2020 | 13.06.2020 | 150680 | 146469 | +2.88 |
| 29.05.2020 | 12.06.2020 | 149620 | 142814 | +4.77 |
| 28.05.2020 | 11.06.2020 | 144630 | 138117 | +4.71 |
| 27.05.2020 | 10.06.2020 | 140540 | 133594 | +5.20 |
| 26.05.2020 | 9.06.2020 | 133350 | 129519 | +2.96 |
| 25.05.2020 | 8.06.2020 | 124460 | 126425 | -1.55 |
| 24.05.2020 | 7.06.2020 | 115230 | 120995 | -4.76 |
| 23.05.2020 | 6.06.2020 | 108420 | 116317 | -6.79 |
| 22.05.2020 | 5.06.2020<sup>17</sup> | 101750 | 111901 | -9.07 |
| 21.05.2020 | 4.06.2020 | 95680 | 106718 | -10.34 |
| 20.05.2020 | 3.06.2020 | 90980 | 101077 | -9.99 |
| 19.05.2020 | 2.06.2020 | 89210 | 97018 | -8.05 |
| 18.05.2020 | 1.06.2020 | 89190 | 93378 | -4.48 |

**Table 8:** Validation of active case prediction for 21 days window

| Date during pre-<br>diction | Date of prediction | Prediction | Actual | Error from<br>actual(%) |
| --- | --- | --- | --- | --- |
| 12.06.2020 | 03.07.2020 | 281980 | 228936 | +23.17 |
| 11.06.2020 | 02.07.2020 | 275210 | 227366 | +21.04 |
| 10.06.2020 | 01.07.2020 | 269210 | 220439 | +22.12 |
| 9.06.2020 | 30.06.2020 | 262650 | 215254 | +22.01 |
| 8.06.2020 | 29.06.2020 | 249590 | 210829 | +18.39 |
| 7.06.2020 | 28.06.2020 | 234960 | 203234 | +15.61 |
| 6.06.2020 | 27.06.2020 | 219520 | 197735 | +11.01 |
| 5.06.2020 | 26.06.2020 | 202420 | 190107 | +6.47 |
| 4.06.2020 | 25.06.2020 | 179170 | 186286 | -3.82 |
| 3.06.2020 | 24.06.2020 | 165570 | 182931 | -9.49 |
| 2.06.2020 | 23.06.2020 | 159800 | 178205 | -10.33 |
| 1.06.2020 | 22.06.2020 | 159190 | 175836 | -9.47 |
| 31.05.2020 | 21.06.2020 | 167410 | 170186 | -1.63 |
| 30.05.2020 | 20.06.2020 | 189100 | 168550 | +12.19 |
| 29.05.2020 | 19.06.2020 | 190740 | 163305 | +16.80 |
| 28.05.2020 | 18.06.2020 | 185060 | 160560 | +15.26 |
| 27.05.2020 | 17.06.2020 | 180950 | 154683 | +16.98 |
| 26.05.2020 | 16.06.2020 | 170900 | 152827 | +11.83 |
| 25.05.2020 | 15.06.2020 | 157520 | 153830 | +2.40 |
| 24.05.2020 | 14.06.2020 | 143090 | 150107 | -4.67 |
| 23.05.2020 | 13.06.2020 | 132980 | 146469 | -9.21 |
| 22.05.2020 | 12.06.2020 | 122580 | 142814 | -14.17 |
| 21.05.2020 | 11.06.2020 | 112990 | 138117 | -18.19 |
| 20.05.2020 | 10.06.2020 | 106000 | 133594 | -20.66 |
| 19.05.2020 | 9.06.2020 | 104500 | 129519 | -19.32 |
| 18.05.2020 | 8.06.2020 | 106360 | 126425 | -15.87 |
| 17.05.2020 | 7.06.2020 | 115190 | 120995 | -4.80 |
| 16.05.2020 | 6.06.2020 | 122680 | 116317 | +5.47 |
| 15.05.2020 | 5.06.2020<sup>18</sup> | 124070 | 111901 | +10.87 |
| 14.05.2020 | 4.06.2020 | 128170 | 106718 | +20.10 |
| 13.05.2020 | 3.06.2020 | 131000 | 101077 | +29.60 |
| 12.05.2020 | 2.06.2020 | 130930 | 97018 | +34.95 |
| 11.05.2020 | 1.06.2020 | 125400 | 93378 | +34.29 |

**Table 9:** Validation of active case prediction for 28 days window

| Date during pre-<br>diction | Date of prediction | Prediction | Actual | Error from<br>actual(%) |
| --- | --- | --- | --- | --- |
| 5.06.2020 | 03.07.2020 | 244420 | 228936 | +6.76 |
| 4.06.2020 | 02.07.2020 | 208060 | 227366 | -8.49 |
| 3.06.2020 | 01.07.2020 | 187420 | 220439 | -14.98 |
| 2.06.2020 | 30.06.2020 | 179740 | 215254 | -16.50 |
| 1.06.2020 | 29.06.2020 | 180490 | 210829 | -14.39 |
| 31.05.2020 | 28.06.2020 | 196170 | 203234 | -3.47 |
| 30.05.2020 | 27.06.2020 | 234880 | 197735 | +18.78 |
| 29.05.2020 | 26.06.2020 | 240880 | 190107 | +26.71 |
| 28.05.2020 | 25.06.2020 | 234600 | 186286 | +25.93 |
| 27.05.2020 | 24.06.2020 | 230870 | 182931 | +26.20 |
| 26.05.2020 | 23.06.2020 | 217000 | 178205 | +21.77 |
| 25.06.2020 | 22.06.2020 | 197330 | 175836 | +12.22 |
| 24.05.2020 | 21.06.2020 | 175470 | 170186 | +3.11 |
| 23.05.2020 | 20.06.2020 | 160680 | 168550 | -4.67 |
| 22.05.2020 | 19.06.2020 | 144820 | 163305 | -11.32 |
| 21.05.2020 | 18.06.2020 | 130030 | 160560 | -19.01 |
| 20.05.2020 | 17.06.2020 | 119630 | 154683 | -22.66 |
| 19.05.2020 | 16.06.2020 | 118580 | 152827 | -22.41 |
| 18.05.2020 | 15.06.2020 | 123330 | 153830 | -19.83 |
| 17.05.2020 | 14.06.2020 | 140410 | 150107 | -6.46 |
| 16.05.2020 | 13.06.2020 | 155690 | 146469 | +6.29 |
| 15.05.2020 | 12.06.2020 | 160390 | 142814 | +12.31 |
| 14.05.2020 | 11.06.2020 | 170060 | 138117 | +23.13 |
| 13.05.2020 | 10.06.2020 | 177650 | 133594 | +32.98 |
| 12.05.2020 | 9.06.2020 | 180350 | 129519 | +39.25 |
| 11.05.2020 | 8.06.2020 | 173260 | 126425 | +37.05 |
| 10.05.2020 | 7.06.2020 | 176620 | 120995 | +45.97 |
| 9.05.2020 | 6.06.2020 | 176260 | 116317 | +51.54 |
| 8.05.2020 | 5.06.2020<sup>19</sup> | 171530 | 111901 | - 53.28 |
| 7.05.2020 | 4.06.2020 | 155630 | 106718 | +45.83 |
| 6.05.2020 | 3.06.2020 | 135500 | 101077 | +34.05 |
| 5.05.2020 | 2.06.2020 | 100131 | 97018 | +3.21 |
| 4.05.2020 | 1.06.2020 | 69995 | 93378 | -25.04 |

**Figure 6:**
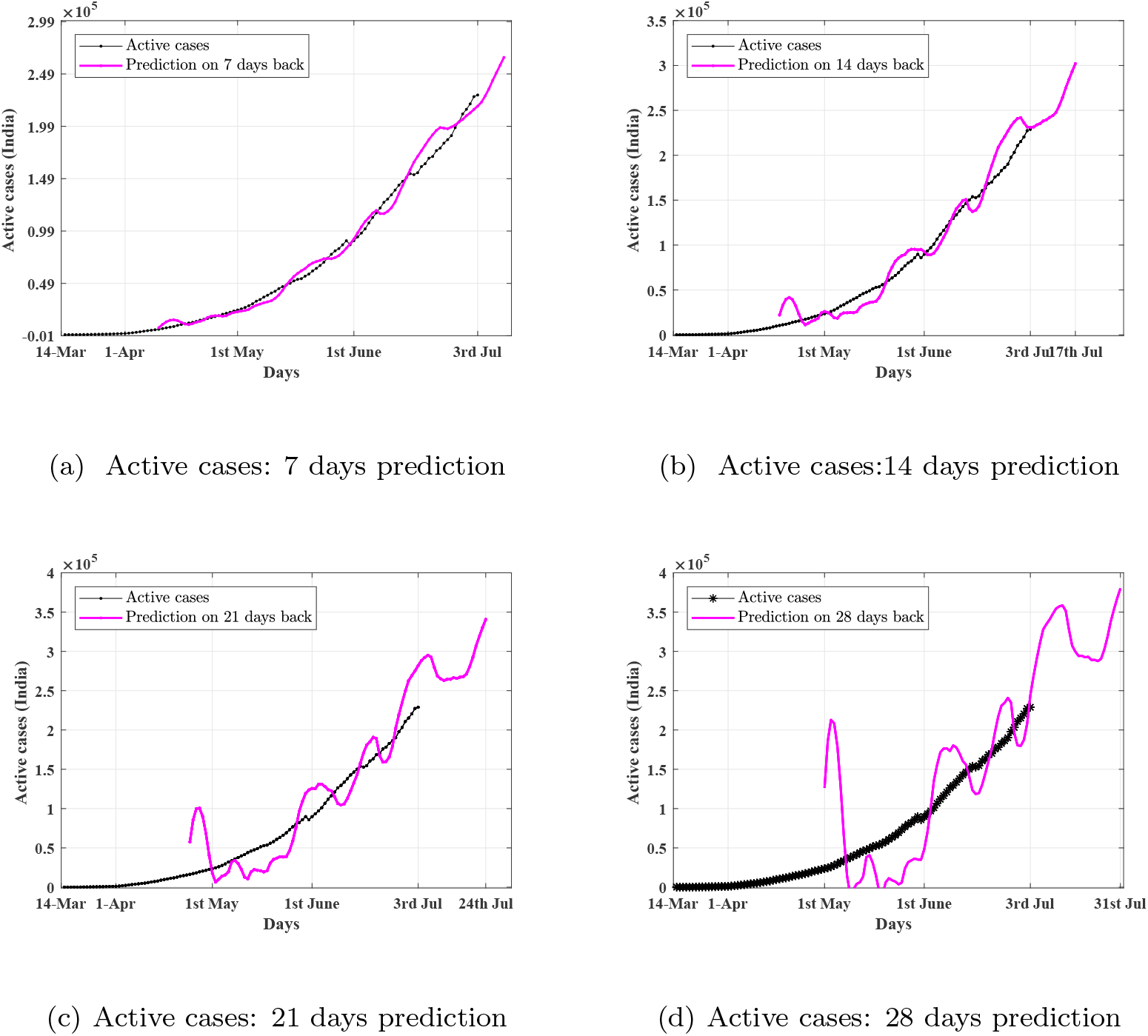
Validation of prediction of active cases

## 6. Discussions

The prediction is good in the smaller time window and error between the actual and predicted case grows with larger time window prediction. The prediction error from June 1^st^ to 3^rd^ July is tabulated in Table 10. The prediction is better in case of total cases compared to active cases. The reason behind the higher value of error bound could be the frequent change in discharge policy by the various state governments to adjust the growth of active patients in hospitals, which in turn affected the recovery rates. Also sometimes the deceased cases are adjusted in a single day after detailed accounting which caused a high jump in the deceased curve. Discharge policy by the governments and reporting of deceased cases have caused the large variation in the actual value of active cases over daily-basis. However, the reporting of the total case is smooth (Figure 3a); therefore, the bound of prediction error is also less.

**Table 10:** Summary of prediction error

| Parameter | 7 days | 14 days | 21 days | 28 days |
| --- | --- | --- | --- | --- |
| Total cases | 0.73 % ± 1.97 % | 1.92 % ± 2.95 % | 4.34 % ± 3.91 % | 6.40 % ± 9.26 % |
| Active cases | 1.24 % ± 6.57 % | 3.04 % ± 10.00 % | 6.33 % ± 16.12 % | 10.20 % ± 24.14 % |

## 7. Future prediction

Future predictions of total cases of India from the proposed algorithm over different time windows are provided in Table 11, Table 12, Table 13, and Table 14. We will compare this table in future to further validate our algorithm. Similar predictions for the active cases are tabulated in Table 15, Table 16, Table 17, and Table 18. The prediction for next 28 days based on the upto date observations are also included in Table 19 for reference.

**Table 11:** Future Prediction of total cases for 7 days window

| Date during prediction | Date of prediction | Prediction | Actual | Error(%) |
| --- | --- | --- | --- | --- |
| 7.06.2020 | 14.07.2020 | 923271 |  |  |
| 8.06.2020 | 15.07.2020 | 953582 |  |  |
| 9.06.2020 | 16.07.2020 | 984856 |  |  |
| 10.06.2020 | 17.07.2020 | 1017049 |  |  |
| 11.07.2020 | 18.07.2020 | 1050612 |  |  |
| 12.07.2020 | 19.07.2020 | 1085283 |  |  |
| 13.07.2020 | 20.07.2020 | 1121190 |  |  |

**Table 12:** Future prediction of total cases for 14 days window

| Date during prediction | Date of prediction | Prediction | Actual | Error(%) |
| --- | --- | --- | --- | --- |
| 30.06.2020 | 14.07.2020 | 904382 |  |  |
| 01.07.2020 | 15.07.2020 | 933350 |  |  |
| 02.07.2020 | 16.07.2020 | 962644 |  |  |
| 03.07.2020 | 17.07.2020 | 994203 |  |  |
| 04.07.2020 | 18.07.2020 | 1028181 |  |  |
| 05.07.2020 | 19.07.2020 | 1064689 |  |  |
| 06.07.2020 | 20.07.2020 | 1102150 |  |  |
| 07.07.2020 | 21.07.2020 | 1138688 |  |  |
| 08.07.2020 | 22.07.2020 | 1175003 |  |  |
| 09.07.2020 | 23.07.2020 | 1212579 |  |  |
| 10.07.2020 | 24.07.2020 | 1251346 |  |  |
| 11.07.2020 | 25.07.2020 | 1292120 |  |  |
| 12.07.2020 | 26.07.2020 | 1334355 |  |  |
| 13.07.2020 | 27.07.2020 | 1378048 |  |  |

**Table 13:** Future prediction of total cases for 21 days window

| Date during prediction | Date of prediction | Prediction | Actual | Error(%) |
| --- | --- | --- | --- | --- |
| 23.06.2020 | 14.07.2020 | 867245 |  |  |
| 24.06.2020 | 15.07.2020 | 893187 |  |  |
| 25.06.2020 | 16.07.2020 | 922109 |  |  |
| 26.06.2020 | 17.07.2020 | 954784 |  |  |
| 27.06.2020 | 18.07.2020 | 990776 |  |  |
| 28.06.2020 | 19.07.2020 | 1030799 |  |  |
| 29.06.2020 | 20.07.2020 | 1070675 |  |  |
| 30.06.2020 | 21.07.2020 | 1107524 |  |  |
| 01.07.2020 | 22.07.2020 | 1141728 |  |  |
| 02.07.2020 | 23.07.2020 | 1176221 |  |  |
| 03.07.2020 | 24.07.2020 | 1214099 |  |  |
| 04.07.2020 | 25.07.2020 | 1255691 |  |  |
| 05.07.2020 | 26.07.2020 | 1301005 |  |  |
| 06.07.2020 | 27.07.2020 | 1347265 |  |  |
| 07.07.2020 | 28.07.2020 | 1391397 |  |  |
| 08.07.2020 | 29.07.2020 | 1434624 |  |  |
| 09.07.2020 | 30.07.2020 | 1479500 |  |  |
| 10.07.2020 | 31.07.2020 | 1525916 |  |  |
| 11.07.2020 | 01.08.2020 | 1575199 |  |  |
| 12.07.2020 | 02.08.2020 | 1626391 |  |  |
| 13.07.2020 | 03.08.2020 | 1679281 |  |  |

**Table 14:** Future prediction of total cases for 28 days window

| **Date during Prediction** | **Date of prediction** | **Prediction** | **Actual** | **Error(%)** |
| --- | --- | --- | --- | --- |
| 16.06.2020 | 14.07.2020 | 875120 |  |  |
| 17.06.2020 | 15.07.2020 | 883710 |  |  |
| 18.06.2020 | 16.07.2020 | 898890 |  |  |
| 19.06.2020 | 17.07.2020 | 920890 |  |  |
| 20.06.2020 | 18.07.2020 | 950760 |  |  |
| 21.06.2020 | 19.07.2020 | 987170 |  |  |
| 22.06.2020 | 20.07.2020 | 1021400 |  |  |
| 23.06.2020 | 21.07.2020 | 1049718 |  |  |
| 24.06.2020 | 22.07.2020 | 1079705 |  |  |
| 25.06.2020 | 23.07.2020 | 1114064 |  |  |
| 26.06.2020 | 24.07.2020 | 1153961 |  |  |
| 27.06.2020 | 25.07.2020 | 1198759 |  |  |
| 28.06.2020 | 26.07.2020 | 1249335 |  |  |
| 29.06.2020 | 27.07.2020 | 1298983 |  |  |
| 30.06.2020 | 28.07.2020 | 1343492 |  |  |
| 01.07.2020 | 29.07.2020 | 1383608 |  |  |
| 02.07.2020 | 30.07.2020 | 1423955 |  |  |
| 03.07.2020 | 31.07.2020 | 1469184 |  |  |
| 04.07.2020 | 01.08.2020 | 1519869 |  |  |
| 05.07.2020 | 02.08.2020 | 1575846 |  |  |
| 06.07.2020 | 03.08.2020 | 1632683 |  |  |
| 07.07.2020 | 04.08.2020 | 1685699 |  |  |
| 08.07.2020 | 05.08.2020 | 1736842 |  |  |
| 09.07.2020 | 06.08.2020 | 1790127 |  |  |
| 10.07.2020 | 07.08.2020 | 1845392 |  |  |
| 11.07.2020 | 08.08.2020 | 1904650 |  |  |
| 12.07.2020 | 09.08.2020 | 1966379 |  |  |
| 13.07.2020 | 10.08.2020 | 2030057 |  |  |

**Table 15:** Future prediction of active cases for 7 days window

| **Date during prediction** | **Date of prediction** | **Prediction** | **Actual** | **Error(%)** |
| --- | --- | --- | --- | --- |
| 7.06.2020 | 14.07.2020 | 305252 |  |  |
| 8.06.2020 | 15.07.2020 | 314019 |  |  |
| 9.06.2020 | 16.07.2020 | 322349 |  |  |
| 10.06.2020 | 17.07.2020 | 330195 |  |  |
| 11.07.2020 | 18.07.2020 | 338128 |  |  |
| 12.07.2020 | 19.07.2020 | 346743 |  |  |
| 13.07.2020 | 20.07.2020 | 357261 |  |  |

**Table 16:** Future prediction of active cases for 14 days window

| **Date during prediction** | **Date of prediction** | **Prediction** | **Actual** | **Error(%)** |
| --- | --- | --- | --- | --- |
| 30.06.2020 | 14.07.2020 | 284328 |  |  |
| 01.07.2020 | 15.07.2020 | 293387 |  |  |
| 02.07.2020 | 16.07.2020 | 302079 |  |  |
| 03.07.2020 | 17.07.2020 | 307619 |  |  |
| 04.07.2020 | 18.07.2020 | 316349 |  |  |
| 05.07.2020 | 19.07.2020 | 327999 |  |  |
| 06.07.2020 | 20.07.2020 | 340698 |  |  |
| 07.07.2020 | 21.07.2020 | 352954 |  |  |
| 08.07.2020 | 22.07.2020 | 363963 |  |  |
| 09.07.2020 | 23.07.2020 | 373654 |  |  |
| 10.07.2020 | 24.07.2020 | 382548 |  |  |
| 11.07.2020 | 25.07.2020 | 391734 |  |  |
| 12.07.2020 | 26.07.2020 | 402159 |  |  |
| 13.07.2020 | 27.07.2020 | 415782 |  |  |

**Table 17:** Future prediction of active cases for 21 days window

| Date during prediction | Date of prediction | Prediction | Actual | Error(%) |
| --- | --- | --- | --- | --- |
| 23.06.2020 | 14.07.2020 | 265641 |  |  |
| 24.06.2020 | 15.07.2020 | 267225 |  |  |
| 25.06.2020 | 16.07.2020 | 267829 |  |  |
| 26.06.2020 | 17.07.2020 | 271197 |  |  |
| 27.06.2020 | 18.07.2020 | 280506 |  |  |
| 28.06.2020 | 19.07.2020 | 292481 |  |  |
| 29.06.2020 | 20.07.2020 | 307021 |  |  |
| 30.06.2020 | 21.07.2020 | 318976 |  |  |
| 01.07.2020 | 22.07.2020 | 330259 |  |  |
| 02.07.2020 | 23.07.2020 | 340672 |  |  |
| 03.07.2020 | 24.07.2020 | 346155 |  |  |
| 04.07.2020 | 25.07.2020 | 356820 |  |  |
| 05.07.2020 | 26.07.2020 | 372312 |  |  |
| 06.07.2020 | 27.07.2020 | 389220 |  |  |
| 07.07.2020 | 28.07.2020 | 405179 |  |  |
| 08.07.2020 | 29.07.2020 | 419071 |  |  |
| 09.07.2020 | 30.07.2020 | 430343 |  |  |
| 10.07.2020 | 31.07.2020 | 440400 |  |  |
| 11.07.2020 | 01.08.2020 | 451062 |  |  |
| 12.07.2020 | 02.08.2020 | 463747 |  |  |
| 13.07.2020 | 03.08.2020 | 481399 |  |  |

**Table 18:** Future prediction of active cases for 28 days window

| **Date during prediction** | **Date of prediction** | **Prediction** | **Actual** | **Error(%)** |
| --- | --- | --- | --- | --- |
| 16.06.2020 | 14.07.2020 | 327825 |  |  |
| 17.06.2020 | 15.07.2020 | 307229 |  |  |
| 18.06.2020 | 16.07.2020 | 299629 |  |  |
| 19.06.2020 | 17.07.2020 | 294172 |  |  |
| 20.06.2020 | 18.07.2020 | 294427 |  |  |
| 21.06.2020 | 19.07.2020 | 292492 |  |  |
| 22.06.2020 | 20.07.2020 | 293131 |  |  |
| 23.06.2020 | 21.07.2020 | 289077 |  |  |
| 24.06.2020 | 22.07.2020 | 289169 |  |  |
| 25.06.2020 | 23.07.2020 | 287667 |  |  |
| 26.06.2020 | 24.07.2020 | 290661 |  |  |
| 27.06.2020 | 25.07.2020 | 302517 |  |  |
| 28.06.2020 | 26.07.2020 | 318523 |  |  |
| 29.06.2020 | 27.07.2020 | 338071 |  |  |
| 30.06.2020 | 28.07.2020 | 353496 |  |  |
| 01.07.2020 | 29.07.2020 | 367618 |  |  |
| 02.07.2020 | 30.07.2020 | 380146 |  |  |
| 03.07.2020 | 31.07.2020 | 385383 |  |  |
| 04.07.2020 | 01.08.2020 | 398498 |  |  |
| 05.07.2020 | 02.08.2020 | 419023 |  |  |
| 06.07.2020 | 03.08.2020 | 441410 |  |  |
| 07.07.2020 | 04.08.2020 | 462128 |  |  |
| 08.07.2020 | 05.08.2020 | 479668 |  |  |
| 09.07.2020 | 06.08.2020 | 492768 |  |  |
| 10.07.2020 | 07.08.2020 | 504113 |  |  |
| 11.07.2020 | 08.08.2020 | 516510 |  |  |
| 12.07.2020 | 09.08.2020 | 531986 |  |  |
| 13.07.2020 | 10.08.2020 | 554767 |  |  |

**Table 19:** Future prediction of active and total cases on 13.07.2020

| **Date of prediction** | **Active case** | **Total case** |
| --- | --- | --- |
| 14.07.2020 | 312279 | 932648 |
| 15.07.2020 | 319461 | 962181 |
| 16.07.2020 | 326767 | 992453 |
| 17.07.2020 | 334198 | 1023476 |
| 18.07.2020 | 341756 | 1055264 |
| 19.07.2020 | 349443 | 1087831 |
| 20.07.2020 | 357261 | 1121190 |
| 21.07.2020 | 365210 | 1155355 |
| 22.07.2020 | 373294 | 1190340 |
| 23.07.2020 | 381513 | 1226157 |
| 24.07.2020 | 389869 | 1262823 |
| 25.07.2020 | 398365 | 1300350 |
| 26.07.2020 | 407002 | 1338754 |
| 27.07.2020 | 415782 | 1378048 |
| 28.07.2020 | 424706 | 1418247 |
| 29.07.2020 | 433777 | 1459366 |
| 30.07.2020 | 442997 | 1501420 |
| 31.07.2020 | 452367 | 1544423 |
| 01.08.2020 | 461890 | 1588391 |
| 02.08.2020 | 471566 | 1633338 |
| 03.08.2020 | 481399 | 1679281 |
| 04.08.2020 | 491390 | 1726234 |
| 05.08.2020 | 501541 | 1774214 |
| 06.08.2020 | 511854 | 1823235 |
| 07.08.2020 | 522331 | 1873314 |
| 08.08.2020 | 532974 | 1924467 |
| 09.08.2020 | 543786 | 1976709 |
| 10.08.2020 | 554767 | 2030057 |

## 8. Conclusions

In this paper, a data-driven prediction model is proposed to predict the total cases and active cases of an area. The model can apply to an area. It is observed that the error between the actual value and predicted value is 4.34 % ± 3.91 % and 6.33 % ± 16.12 % for total cases and active cases respectively over the prediction window of 21 days. We have also provided the prediction for the next 28 days from today for further validation of the proposed algorithm. The short term prediction can be used in the allocation of scar medical resources among different units, optimal lockdown planning.

## Data Availability

All data is available in open source.

## ACKNOWLEDGEMENTS

The authors would like to thank Mayur Shewale and GCDSL lab members for their suggestions in data processing and analysis of results.

## References

[1] C. Zhan, C. K. Tse, Z. Lai, T. Hao, J. Su, Prediction of covid-19 spreading profiles in south korea, italy and iran by data-driven coding, medRxiv.

[2] S. Kim, Y. B. Seo, E. Jung, Prediction of covid-19 transmission dynam- ics using a mathematical model considering behavior changes in korea., Epidemiology and Health 42 (2020) 1–6.

[3] M. Mandal, S. Jana, S. K. Nandi, A. Khatua, S. Adak, T. Kar, A model based study on the dynamics of covid-19: Prediction and control., Chaos Solitons and Fractals 136 (2020) 109889–109889.

[4] W. C. Roda, M. B. Varughese, D. Han, M. Y. Li, Why is it difficult to accurately predict the covid-19 epidemic?, Infectious Disease Modelling 5 (5) (2020) 271–281.

[5] A. Rajesh, H. Pai, V. Roy, S. Samanta, S. Ghosh, Covid-19 prediction for india from the existing data and sir(d) model study, medRxiv.

[6] S. Mondal, S. Ghosh, Possibilities of exponential or sigmoid growth of covid19 data in different states of india, medRxiv.

[7] J. Singh, P. K. Ahluwalia, A. Kumar, Mathematical model based covid-19 prediction in india and its different states, medRxiv.

[8] Y. Zhang, C. You, Z. Cai, J. Sun, W. Hu, X.-H. Zhou, Prediction of the covid-19 outbreak based on a realistic stochastic model, medRxiv.

[9] A. J. Kucharski, T. W. Russell, C. Diamond, Y. Liu, J. Edmunds, S. Funk, R. M. Eggo, F. Sun, M. Jit, J. D. Munday, N. Davies, A. Gimma, K. van Zandvoort, H. Gibbs, J. Hellewell, C. I. Jarvis, S. Clifford, B. J. Quilty, N. I. Bosse, S. Abbott, P. Klepac, S. Flasche, Early dynamics of transmission and control of covid-19: a mathematical modelling study., Lancet Infectious Diseases 20 (5) (2020) 553–558.

[10] L. Peng, W. Yang, D. Zhang, C. Zhuge, L. Hong, Epidemic analysis of covid-19 in china by dynamical modeling., arXiv preprint 2002.06563.

[11] B. Zareie, A. Roshani, M. A. Mansournia, M. A. Rasouli, G. Moradi, A model for covid-19 prediction in iran based on china parameters., Archives of Iranian Medicine 23 (4) (2020) 244–248.

[12] J. D. Annan, J. C. Hargreaves, Model calibration, nowcasting, and operational prediction of the covid-19 pandemic, medRxiv.

[13] G. Barmparis, G. Tsironis, Estimating the infection horizon of covid-19 in eight countries with a data-driven approach, Chaos, Solitons and Fractals (2020) 109842.

[14] S. Sanchez-Caballero, M. A. Selles, M. A. Peydro, E. Perez-Bernabeu, An efficient covid-19 prediction model validated with the cases of china, italy and spain: Total or partial lockdowns?, Journal of Clinical Medicine 9 (5) (2020) 1547–1547.

[15] A. B. Younes, Z. Hasan, Covid-19: Modeling, prediction, and control, Ap- plied Sciences 10 (11) (2020) 3666.

[16] M. C. Sabat, S. A. Muoz, E. lvarez Lacalle, D. L. Codina, P. J. C. Iglesias, C. P. Soler, Empiric model for short-time prediction of covid-19 spreading, PLOS Computational Biology (2020) 1–14.

[17] S. Gupta, R. Shankar, Estimating the number of covid-19 infections in indian hot-spots using fatality data, arXiv preprint 2004.04025.

[18] D. K. K. Singh, S. Kumar, P. Dixit, D. M. K. Bajpai, Kalman filter based short term prediction model for covid-19 spread, medRxiv.

[19] J.-P. Quadrat, 1-c nonlinear covid-19 epidemic model and application to the epidemic prediction in france, medRxiv.

[20] A. Tomar, N. Gupta, Prediction for the spread of covid-19 in india and effectiveness of preventive measures, Science of The Total Environment (2020) 138762.

[21] S. Singh, K. S. Parmar, J. Kumar, S. J. S. Makkhan, Development of new hybrid model of discrete wavelet decomposition and autoregressive integrated moving average (arima) models in application to one month forecast the casualties cases of covid-19., Chaos Solitons and Fractals 135 (2020) 109866.

[22] S. I. Alzahrani, I. A. Aljamaan, E. A. Al-Fakih, Forecasting the spread of the covid-19 pandemic in saudi arabia using arima prediction model under current public health interventions., Journal of Infection and Public Health.

